# Distance- and Hierarchy-Dependent Functional Dysconnectivity in Schizophrenia and Its Association with Cortical Microstructure

**DOI:** 10.1101/2025.07.28.25332321

**Authors:** Isaac David, Shuntaro Sasai, Felipe Branco de Paiva, Melanie Boly, Giulio Tononi, Larissa Albantakis

## Abstract

**Background:** Schizophrenia is associated with widespread functional dysconnectivity, but the spatial scale and structural correlates of these alterations remain unclear. Short-range connectivity, in particular, has received limited attention due to methodological constraints, despite its relevance to local microcircuit dysfunction.

**Methods:** We applied a vertex-wise, distance-dependent analysis of functional connectivity strength (FCS) to resting-state fMRI data from 86 schizophrenia patients and 99 healthy controls across two datasets. FCS was partitioned by geodesic distance on the cortical surface and analyzed by cortical hierarchy. We also assessed two proxies of intracortical microstructure: T1w/T2w ratio and a novel signal-detection-based measure of individualized data-driven functional connectivity density (idFCD).

**Results:** Schizophrenia patients exhibited reductions in short-range FCS within the dorsal primary somatosensory cortex. These functional alterations colocalized with abnormalities in both microstructural proxies and were not evident in global FCS analysis. In contrast, longer-range FCS was increased in transmodal regions, particularly the precuneus, without associated microstructural differences. Hierarchical analysis confirmed this dissociation, with structure– function disruption in primary networks and increased relative FCS in transmodal regions without microstructural association.

**Conclusions:** Our findings support two distinct patterns of cortical dysconnectivity in schizophrenia: short-range reductions in primary sensory areas that colocalize with microstructural abnormalities, and longer-range increases in transmodal regions that appear structurally decoupled at the local level. By integrating distance-dependent functional measures with independent proxies of intracortical microstructure, this study highlights the underappreciated role of short-range connectivity disruptions in primary areas and provides a complementary framework to conventional approaches based on regional or global analyses and diffusion-weighted imaging.

## 1. Introduction

Schizophrenia is associated with widespread disruptions in brain connectivity, yet its underlying mechanisms remain unclear. The dysconnectivity hypothesis posits that the disorder’s cognitive and clinical symptoms arise from abnormal functional and structural connectivity (Friston and Frith 1995; Stephan et al. 2009), leading to deficits in both information convergence and divergence (Tononi and Edelman 2000; Lord et al. 2017). These disruptions have been linked to altered thalamocortical communication (Ferrarelli and Tononi 2011; Giraldo-Chica and Woodward 2017; Baran et al. 2019), impaired activity flow (Hearne et al. 2021), and widespread changes in synaptic plasticity and network topology (Bullmore et al. 1997; Stephan et al. 2006; Lynall et al. 2010). Despite extensive research, a clear and unified account of schizophrenia in terms of aberrant structural and functional connectivity remains elusive but critically necessary to elucidate underlying disease mechanisms.

Two central challenges in characterizing dysconnectivity in schizophrenia are (1) reconciling structural and functional connectivity findings (van den Heuvel and Fornito 2014; Gao et al. 2023; W. Zhao et al. 2020) and (2) determining the relative contributions of different brain networks to the disorder (Gong et al. 2020; D. Dong et al. 2023; Holmes et al. 2023). While structural imaging studies tend to report atrophy and hypoconnectivity, functional neuroimaging findings have been more variable, with inconsistent reports of both hyper- and hypoconnectivity across different networks (Pettersson-Yeo et al. 2011; Fornito et al. 2012; Fornito and Bullmore 2015; Ding et al. 2019; L. Guo et al. 2023). Moreover, studies have found little colocalization between structural and functional abnormalities (W. Zhao et al. 2020).

One possible explanation is the limited ability of diffusion-weighted magnetic resonance imaging (dw-MRI) to capture local and short-range cortico-cortical connections, which together account for over 80% of all synapses in sensory cortex (Markov et al. 2011). Axonal projections within sensory areas seldom extend beyond 5 mm (Stettler et al. 2002), and exhibit isotropic diffusion along the pial surface due to the grid-like lateral organization of neurites (Shimony et al. 1999; Jespersen et al. 2012). Beyond strictly local connectivity, the density of feedforward and feedback connections to sensory cortex usually peaks at neighboring regions (Markov et al. 2011). As a result, structural connectivity (SC) at short-ranges remains largely “intractable” by standard white-matter tractography (but see Jespersen et al. (2012) and and H. Zhang et al. (2012)). Importantly, local microstructure contributes to neural processing (Angelucci and Bressloff 2006; Lee et al. 2012; H. Liang et al. 2017; Davis et al. 2020), and disease-related abnormalities in short-range SC may account in part for mesoscopic changes in functional connectivity (FC) in schizophrenia (Must et al. 2004; Fornito et al. 2012). However, standard FC analyses rely on parcellation-based methods, which introduce coarse-graining that can obscure finer-grained local dysconnectivity. Nevertheless, emerging evidence suggests that short-range connectivity alterations may contribute to schizophrenia (C. Zhao et al. 2018; Yu et al. 2021; Cai et al. 2022).

With respect to the relative contributions of different brain networks to the pathophysiology of schizophrenia, cortical organization follows a principal macroscale gradient from unimodal (sensorimotor, visual) to transmodal (default mode, frontoparietal) networks, reflecting increasing abstraction of information (Margulies et al. 2016). Notably, this hierarchy is spatially embedded, with transmodal regions situated farthest from unimodal sensorimotor areas along the cortical surface. Early studies primarily implicated disruptions in frontoparietal and frontotemporal circuits, indicating top-down dysregulation in schizophrenia (Minzenberg et al. 2009; Ellison-Wright and Bullmore 2009; Fornito et al. 2012; Dima et al. 2010). However, more recent evidence suggests that dysconnectivity is not confined to higher-order networks but also affects sensory and motor systems (Yoon et al. 2010; Dondé et al. 2019; Bortolon et al. 2015). Alterations in somatomotor, visual and auditory networks have been observed alongside disruptions in default mode, ventral attention (salience), and frontoparietal (executive) networks (D. Dong et al. 2018; Li et al. 2019; Gong et al. 2020). Taken together, these findings underscore the need for a more systematic investigation of how dysconnectivity varies across spatial scales and cortical hierarchies.

Here, we investigated distance-dependent alterations in cortical functional connectivity strength (FCS) in schizophrenia and examined their relationship to both macroscale network hierarchy and inferred microstructural abnormalities. Using resting-state fMRI data from two available datasets (86 schizophrenia patients, 99 healthy controls total), we derived vertex-wise cortico-cortical FC in a surface-based representation at multiple spatial scales and grouped into primary, intermediate, and transmodal networks. To bridge functional and structural findings, we further analyzed two proxy measures of microstructural connectivity: T1/T2 ratios (a marker of gray matter integrity) and functional connectivity density (FCD), thresholded using an anatomically constrained approach to minimize false positives.

Short-range FCS deficits were predominantly observed in sensory networks, particularly the somatosensory cortex, and were accompanied by colocalized microstructural abnormalities in the primary somatomotor cortex. These functional and structural alterations were further supported by statistical correlations that varied as a function of both distance and network hierarchy. In contrast, long-range FCS increases were identified in higher-order transmodal networks, particularly the precuneus, but only when considering relative (z-scored) FCS, with no corresponding microstructural disruption. Together, these analyses provide a multi-level characterization of dysconnectivity in schizophrenia, offering new insights into the relationship between short- and long-range dysconnectivity and their cortical organization.

## 2. Materials and methods

### 2.1 Data

We used two independent resting-state fMRI (rs-fMRI) datasets from the SchizConnect repository (L. Wang et al. 2016): the Center for Biomedical Research Excellence (COBRE) and the Northwestern University Neuroimaging Data Archive (NUNDA/NMorphCH) datasets (Aine et al. 2017; Alpert et al. 2016). The COBRE dataset remains available through the International Neuroimaging Data-sharing Initiative (INDI) via NITRC (fcon_1000.projects.nitrc.org/indi/retro/cobre.html). Healthy controls (HC) and patients with schizophrenia spectrum disorder (SSD) were sourced from both datasets. SSD encompasses a range of mental health conditions characterized by delusions, hallucinations, disorganized thinking and abnormal behavior. We considered schizophrenia proper and schizoaffective disorder, which are diagnosed differentially only by heightened concomitant mood disorder symptoms in the case of schizoaffective disorder. Only baseline recordings were included for analysis. BOLD resting-state fMRI (rs-fMRI) recordings were used to calculate distance-dependent functional connectivity and individualized data-driven functional connectivity density (idFCD) (see below). T1/T2 integrity was assessed using the ratio of T1w and T2w image modalities of brain anatomy. Screening for missing image modalities (BOLD, T1w, T2w) resulted in the dismissal of 67 COBRE subjects. Motion-related exclusion was applied using a framewise displacement threshold of 0.5 mm to define outlier time points (Power et al. 2012); subjects with more than 50% of frames classified as motion outliers were excluded. This resulted in the removal of 12 patients (11 COBRE, 1 NMorphCH) and 5 controls (4 COBRE, 1 NmorphCH). The final sample included 86 SSD patients (43 COBRE, 43 NMorphCH, 82 schizophrenia, 4 schizoaffective disorder) and 99 controls (59 COBRE, 40 NMorphCH), with mean age (and standard deviation) of 34.27 (10.44) and 33.47 (10.51) years, respectively (see Table 2). All experimental procedures were approved by local ethics committees, and written informed consent was obtained from all participants in the original studies.

### 2.2 Imaging parameters

Both datasets were acquired using 3T Siemens scanners. The COBRE rs-fMRI sequence consisted of 150 time points (5 min at TR = 2 s). For NMorphCH, two runs from the same session were concatenated, resulting in 328 time points per subject (12.026 min at TR = 2.2 s, or 13.666 min at TR = 2.5 s). Sequence parameters of all three image modalities are detailed in Table 1.

**Table 1:** Sequence parameters used for the MRI protocols.

|  | COBRE |  |  | NMorphCH |  |  |
| --- | --- | --- | --- | --- | --- | --- |
|  | fMRI | T1w | T2w | fMRI | T1w | T2w |
| **TR (ms)** | 2000 | 2530 | 15500 | 2200, 2500 | 2400 | 3200 |
| **TE (ms)** | 29 | 1.64, 3.5, 5.36, 7.22, 9.08 (multi-echo) | 77 | 20, 27 | 3.16 | 455 |
| **TI (ms)** |  | 900 |  |  |  |  |
| **Flip angle** | 75° | 7° | 155° | 90° | 8° | 120° |
| **Voxel dimensions (mm)** | 3.75 × 3.75 × 4.55 | 1 × 1 × 1 | 1.1 × 1.1 × 1.5 | 3 × 3 × 3, 4 × 4 × 4 | 1 × 1 × 1 | 1 × 1 × 1 |
| **Slices** | 32 | 176 | 120 | 36 | 176 | 176 |
| **Slice resolution** | 64 × 64 | 256 × 256 | 192 × 192 | 64 × 64 | 256 × 256 | 256 × 256 |

### 2.3 Data preprocessing

All preprocessing was performed using *fMRIPrep* 23.1.4 ((Esteban et al. 2019; Markiewicz et al. 2024) RRID:SCR_016216), based on *Nipype* 1.8.6 ((Gorgolewski et al. 2011; Esteban et al. 2022) RRID:SCR_002502). *fMRIPrep* automatically generates detailed reports of the preprocessing steps applied, which are included below.

#### 2.3.1 Anatomical data preprocessing

For each subject, the T1-weighted (T1w) image was corrected for intensity non-uniformity (INU) with *N4BiasFieldCorrection* (Tustison et al. 2010), distributed with ANTs ((Avants et al. 2008) RRID:SCR_004757), and used as T1w-reference throughout the workflow. The T1w-reference was then skull-stripped with a *Nipype* implementation of the *antsBrainExtraction.sh* workflow (from ANTs), using OASIS30ANTs as target template. Brain tissue segmentation of cerebrospinal fluid (CSF), white-matter (WM) and gray-matter (GM) was performed on the brain-extracted T1w using *fast* (FSL, RRID:SCR_002823, (Y. Zhang et al. 2001)). Brain surfaces were reconstructed using recon-all (FreeSurfer 7.3.2, RRID:SCR_001847, (Dale et al. 1999)), and the brain mask estimated previously was refined with a custom variation of the method to reconcile ANTs-derived and FreeSurfer-derived segmentations of the cortical gray-matter of Mindboggle (RRID:SCR_002438, (Klein et al. 2017)). *Grayordinate* “dscalar” files (Glasser et al. 2013) containing 91k samples were also generated using the highest-resolution *fsaverage* as an intermediate standardized surface space. Volume-based spatial normalization to a standard space (MNI152NLin2009cAsym) was performed through nonlinear registration with *antsRegistration* (ANTs), using brain-extracted versions of both T1w reference and the T1w template. The following templates were selected for spatial normalization and accessed with *TemplateFlow* (23.0.0, (Ciric et al. 2022)): *ICBM 152 Nonlinear Asymmetrical template version 2009c* [(Fonov et al. 2009), RRID:SCR_008796; TemplateFlow ID: MNI152NLin2009cAsym].

#### 2.3.2 Functional data preprocessing

For each of the BOLD runs found per subject, the following preprocessing was performed. First, a reference volume and its skull-stripped version were generated using a custom methodology of *fMRIPrep*. Head-motion parameters with respect to the BOLD reference (transformation matrices, and six corresponding rotation and translation parameters) are estimated before any spatiotemporal filtering using *mcflirt* (FSL, (Jenkinson et al. 2002)). BOLD runs were slice-time corrected to 0.974s (0.5 of slice acquisition range 0s-1.95s) using *3dTshift* from AFNI ((Cox and Hyde 1997), RRID:SCR_005927). The BOLD time-series (including slice-timing correction when applied) were resampled onto their original, native space by applying the transforms to correct for head-motion. These resampled BOLD time-series will be referred to as *preprocessed BOLD in original space*, or just *preprocessed BOLD*. The BOLD reference was then co-registered to the T1w reference using *bbregister* (FreeSurfer) which implements boundary-based registration (Greve and Fischl 2009). Co-registration was configured with six degrees of freedom. Several confounding time-series were calculated based on the *preprocessed BOLD*: framewise displacement (FD), DVARS and three region-wise global signals. FD was computed using two formulations following Power (absolute sum of relative motions, (Power et al. 2014)) and Jenkinson (relative root mean square displacement between affines, (Jenkinson et al. 2002)). FD and DVARS are calculated for each functional run, both using their implementations in *Nipype* (following the definitions by Power et al. (2014)). The three global signals are extracted within the CSF, the WM, and the whole-brain masks. Additionally, a set of physiological regressors were extracted to allow for component-based noise correction (*CompCor*, (Behzadi et al. 2007)). Principal components are estimated after high-pass filtering the *preprocessed BOLD* time-series (using a discrete cosine filter with 128s cut-off) for the two *CompCor* variants: temporal (tCompCor) and anatomical (aCompCor). tCompCor components are then calculated from the top 2% variable voxels within the brain mask. For aCompCor, three probabilistic masks (CSF, WM and combined CSF+WM) are generated in anatomical space. The implementation differs from that of Behzadi et al. in that instead of eroding the masks by 2 pixels on BOLD space, a mask of pixels that likely contain a volume fraction of GM is subtracted from the aCompCor masks. This mask is obtained by dilating a GM mask extracted from the FreeSurfer’s *aseg* segmentation, and it ensures components are not extracted from voxels containing a minimal fraction of GM. Finally, these masks are resampled into BOLD space and binarized by thresholding at 0.99 (as in the original implementation). Components are also calculated separately within the WM and CSF masks. For each CompCor decomposition, the *k* components with the largest singular values are retained, such that the retained components’ time series are sufficient to explain 50 percent of variance across the nuisance mask (CSF, WM, combined, or temporal). The remaining components are dropped from consideration. The head-motion estimates calculated in the correction step were also placed within the corresponding confounds file. The confound time series derived from head motion estimates and global signals were expanded with the inclusion of temporal derivatives and quadratic terms for each (Satterthwaite et al. 2013). Frames that exceeded a threshold of 0.5 mm FD or 1.5 standardized DVARS were annotated as motion outliers. Additional nuisance timeseries are calculated by means of principal components analysis of the signal found within a thin band (*crown*) of voxels around the edge of the brain, as proposed by (Patriat et al. 2017). The BOLD time-series were resampled into standard space, generating a *preprocessed BOLD run in MNI152NLin2009cAsym space*. First, a reference volume and its skull-stripped version were generated using a custom methodology of *fMRIPrep*. The BOLD time-series were resampled onto the left/right-symmetric template “fsLR” (Glasser et al. 2013). *Grayordinates* files (Glasser et al. 2013) containing 91k samples were also generated using the highest-resolution *fsaverage* as intermediate standardized surface space. All resamplings can be performed with *a single interpolation step* by composing all the pertinent transformations (i.e. head-motion transform matrices, susceptibility distortion correction when available, and co-registrations to anatomical and output spaces). Gridded (volumetric) resamplings were performed using *antsApplyTransforms* (ANTs), configured with Lanczos interpolation to minimize the smoothing effects of other kernels (Lanczos 1964). Non-gridded (surface) resamplings were performed using *mri_vol2surf* (FreeSurfer).

Many internal operations of *fMRIPrep* use *Nilearn* 0.10.1 ((Abraham et al. 2014), RRID:SCR_001362), mostly within the functional processing workflow. For more details of the pipeline, see the section corresponding to workflows in *fMRIPrep*’s documentation.

### 2.4 Functional connectivity (FC)

#### 2.4.1 Additional processing steps

To mitigate motion-related artifacts that could introduce spurious correlations in BOLD signals (Caparelli et al. 2005; Power et al. 2015), nuisance regression was performed using average white matter signal, average CSF signal, and motion regressors computed by fMRIprep. Motion regressors included three translation and three rotation parameters, their first time derivatives, and motion spikes identified via framewise displacement as described above (Power et al. 2012). Linear detrending and bandpass filtering (5th-order Butterworth filter; 0.01– 0.1 Hz) were applied to remove scanner-related thermal drifts and high-frequency physiological noise (Cordes et al. 2001; Foerster et al. 2005).

#### 2.4.2 Vertex-wise distance-dependent functional connectivity

Using Pearson correlation, cortical FC was computed between pairs of surface vertices from individual BOLD surface time series in *fsLR* space. No thresholding was applied, and all correlation values were retained. The full per-subject correlation matrices—one for each cortical hemisphere—were used to derive distance-dependent FC matrices as follows. To assess vertex-to-vertex distances, each hemisphere was inflated into a sphere with radius of 50 mm. Next, for each vertex, only correlations with surface vertices within a specified distance range (measured by concentric circles on the spherical surface centered at that vertex) were retained for analysis. A vertex’s distance-dependent functional connectivity strength (FCS) was calculated as its mean FC with all other vertices within the specified distance range (X. Liang et al. 2013). FCS was evaluated for the following distance ranges (square bracket and parenthesis denote “inclusive” and “exclusive”, respectively): [0 mm to 5 mm), [5 mm to 15 mm), [15 mm to 30 mm), [30 mm to 60 mm), [60 mm to 120 mm), and [120 mm to 250 mm), as well as for the full (unmasked) correlation matrix (“all”). [0 mm to 5 mm) and [5 mm to 15 mm) were split into separate ranges to avoid overlap because the former spans less than 2 individual vertices in any direction and accommodates 95.05% of the probability density of our smoothing kernel, potentially inflating FCS estimates. Both ranges were analyzed identically but treated separately to avoid confounding effects from local smoothing. Distance increases for subsequent ranges roughly follow geometric growth according to the recurrence *r_n_ = r_0_ · 2^n^, r_0_ = 3.75 mm, n*∈*{1, 2, 3, 4, 5}*. Individual distance-dependent FCS maps were normalized using Fisher’s z-transform. We refer to this measure as absolute FCS (aFCS). Additionally, the resulting values were standardized into z-scores relative to each subject’s aFCS distribution, yielding relative FCS (rFCS) values that minimize potential random effects due to individual or dataset variability. For brevity, we will use “FCS” to refer to both aFCS and rFCS jointly. Finally, aFCS and rFCS values were smoothed using a bivariate Gaussian kernel (FWHM = 3 mm).

#### 2.4.3 Macroscale network hierarchy

To further investigate the anatomical distribution and origin of differences in distance-dependent FCS, vertices from the FCS maps were categorized into a three-level network hierarchy, reflecting the topological gradient thought to underlie gross cortical organization (Margulies et al. 2016; Felleman and Van Essen 1991; Mesulam 1998; Huang and Woodward 2023). This gradient spans from primary sensory and motor networks, through intermediate heteromodal attentional networks, to transmodal association networks.

Smoothed and normalized FCS values were first mapped vertex-wise to the resting-state network parcellation by Thomas Yeo et al. (2011). Following Margulies et al. (2016), each network from this parcellation was then assigned to one of three hierarchical categories: primary (visual and somatomotor networks), intermediate (ventral and dorsal attention networks), and transmodal (frontoparietal, limbic and default mode networks). Mean FCS values for each category were then obtained by averaging across the respective vertices.

#### 2.4.4 Individualized data-driven functional connectivity density (idFCD)

In the absence of microstructural cortical data beyond T1w and T2w images, we derived a surrogate measure for a subject’s structural connectivity from their functional connectivity, termed individualized data-driven functional connectivity density (idFCD). This measure was used to generate spatial maps of node degree, a simple graph-theoretic metric of centrality (Rubinov and Sporns 2010). idFCD was calculated as follows: Preprocessed time series were extracted from the cuneus and its contralateral middle frontal gyrus (MFG, rostral portion) in both hemispheres, following the anatomically-defined atlas by Desikan et al. (2006). These regions have no known direct anatomical fasciculus. Vertex-level between-ROI correlations were computed, and each subject’s maximum correlation was used as a threshold for binarizing their original vertex-level FC matrix (non-distance-dependent). This conservative criterion ensures that all FC edges weaker than the strongest (representative) correlation between anatomically unconnected regions were discarded, leaving only edges suggestive of anatomical connections. In other words, our approach minimized type-I errors in detecting pseudo-structural connections, albeit at the expense of true positives. The resulting binarized adjacency matrix was summed along one dimension, yielding a node-degree vector for an undirected, vertex-resolution network. Overall, this approach is conceptually similar to functional connectivity density (FCD) mapping and related methods (van den Heuvel et al. 2009; Buckner et al. 2009; Tomasi and Volkow 2010), but with a data-driven threshold that was anatomically informed rather than arbitrarily chosen. Finally, idFCD maps were z-scored relative to each subject’s distribution and smoothed using a bivariate Gaussian kernel (FWHM = 3 mm) before assessing group differences.

### 2.5 T1/T2

We also examined changes in the so-called “myelin map”, defined as the ratio of raw (i.e. uncorrected for magnetic field bias) T1w to T2w images (Glasser and Van Essen 2011). While histological validations and optical imaging studies suggest that this ratio is not a direct correlate of myelin content, it remains an independent marker of gray matter integrity (Mühlau 2022; Sandrone et al. 2023). T2w images were registered with their T1w homologues in subject-space using FSL’s *flirt* tool with trilinear interpolation (Jenkinson et al. 2002). After computing the T1/T2 ratio, we applied a two-tailed outlier removal, discarding the most extreme 2% of voxels. For cortical surface projection, we followed the same procedure used by *fMRIprep* for resampling BOLD timeseries. Specifically, six equidistant 164k *fsaverage* surfaces were extracted from the cortical ribbon using *FreeSurfer’s* segmentation, with the outermost layers corresponding to the white matter and pial borders. These surfaces were averaged using FreeSurfer’s *mri_vol2surf*. The mean T1/T2 surface in *fsaverage* space was then downsampled to the 32k *fsLR* surface via HCP’s *wb_command* with the metric-resample option, using BARYCENTRIC (i.e. nearest neighbor) interpolation and spherical versions of the source and target manifolds. As with idFCD, the same z-scoring, smoothing, and statistical procedures were followed to measure group differences in T1/T2 integrity.

### 2.6 Statistical analysis

Group differences in vertex-wise analyses (FCS, idFCD, and T1/T2 integrity) were assessed using two-sample z-tests with a cluster-forming threshold of p < 0.001. To control the family-wise error rate (FWE), a permutation test was performed, generating the null distribution of maximum cluster sizes via 10,000 label permutations of the diagnostic groups. Unless otherwise specified, reported clusters in the results section are those that survived an FWE-corrected significance threshold of p < 0.05, given the empirically estimated max-statistic null distribution.

For the macroscale network-hierarchy analysis, we fitted the following linear mixed-effects model (LMM) to mean rFCS and aFCS:

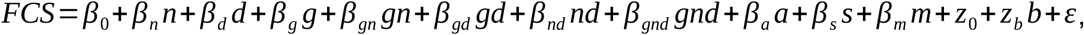

in order to assess the intercept (β0), effects due to network hierarchy (*n*: primary, intermediate, transmodal), distance (*d*: six spatial scales + “all”), diagnostic group (*g*: SSD vs. HC), as well as their interactions, while accounting for potential confounds (age (*a*), sex (*s*) and percentage of motion (*m*) outliers), plus a random intercept (*z_0_*) and linear effect due to data set (*b:* COBRE vs. NMorphCH). Post-hoc pairwise comparisons were conducted to examine SSD vs. HC differences within each {distance range, network-hierarchy level} combination. P-values were estimated nonparametrically from 10,000 permutations of diagnosis labels and corrected for multiple comparisons using the Holm-Bonferroni method.

For correlation analyses between distinct brain measurements, simple linear regression models and Pearson correlations were computed for pairs of z-maps derived from previous group comparisons (ΔFCS, ΔidFCD, and Δ(T1/T2)), also separating by distance and hierarchy level. For every distance, pairwise comparisons of correlations were assessed from differences of mean normalized cross-product deviations, as obtained from a 2-factor ANOVA (3 network levels, 7 distance ranges) plus an interaction term between the 2 factors. Normalized cross-product deviations correspond to the terms that get averaged in the definition of the Pearson correlation. Resulting p-values were again FWE-corrected using the Holm-Bonferroni method.

PANSS scores were available for patients in the COBRE data set only. To investigate the relationship between FCS alterations and symptomatology, we used average rFCS values masked by the anatomical clusters demonstrating significant between-group differences, as well as average rFCS values grouped by hierarchical level and distance. Both were fitted to PANSS scores (positive, negative, and other) using multiple linear regression, from which pairwise correlations and effect-size p-values were obtained. To better characterize symptom structure, we also performed a principal component analysis (PCA) on the covariance matrix of all 30 PANSS scores before fitting the first three components (explaining 89% of variance) with the rFCS measures. In all cases, sex, age, and head-motion were controlled for as extra confounds in the regression models. We highlight correlations associated to parameter estimates with p < 0.05. FWE-correction was attempted, but no correlations survived.

## 3. Results

We compared cortical FCS, idFCD, and T1/T2 integrity between schizophrenia patients and healthy controls. FCS was analyzed across multiple distance ranges to investigate potential alterations in connectivity patterns at finer spatial scales. To assess whether these alterations followed established macroscale gradients by Margulies et al. (2016), we categorized the distance-dependent FCS into primary, intermediate, and transmodal networks. To explore the relationship between microstructural and functional changes, we examined idFCD and T1/T2 alterations, their anatomical colocalization, and statistical associations with FCS differences, both at the vertex level and across macroscale ROIs.

### 3.1 Demographics and clinical information

We analyzed two publicly available datasets (COBRE and NMorphCH). After excluding subjects due to incomplete data and excessive head motion, the final sample included 43 patients and 59 controls from COBRE and 43 patients and 40 controls from NMorphCH. PANSS scores were available for COBRE but not for NMorphCH patients. See Table 2 for detailed demographic information. There was no significant difference in age between groups; however, the schizophrenia group included higher proportions of males and motion outliers than the control group. P-values were obtained using multivariate logistic regression with data set, age, sex, and percentage of motion outliers as independent variables.

**Table 2:** Demographic and clinical characteristics.

| Variable | SSD (n = 86) | Controls (n = 99) | p-value |
| --- | --- | --- | --- |
| Data set<br>(Cobre/NMorphCH) | 43/43 | 59/40 | 0.083 |
| Age<br>(Years; mean (SD)) | 34.27 (10.44) | 33.47 (10.51) | 0.54 |
| Sex (male/female) | 65/21 | 59/40 | 0.028 |
| Motion outliers<br>(%. Mean (SD)) | 32.18 (31.72) | 17.37 (18.62) | 0.0014 |
| PANSS (COBRE)<br>(mean (SD)) | Positive: 17.20 (2.96)<br>Negative: 15.56 (3.96)<br>Other: 27.53 (7.52)<br>Total: 60.30 (10.90) | NA | NA |

### 3.2 Distance-dependent Functional Connectivity Strength

#### 3.2.1 Vertex-wise FCS

Distance-dependent analysis of rFCS revealed significant group differences that were not apparent in global rFCS. Compared to controls, schizophrenia patients exhibited a bilateral short-range functional connectivity deficit in the dorsal primary somatosensory cortex (S1) and increased longer-range connectivity in the dorsoposterior precuneus (Fig. 1). Specifically, S1 showed reduced rFCS at the 0-5 mm (right hemisphere), 5-15 mm (bilaterally) and 15-30 mm ranges (bilaterally), while the precuneus exhibited increased connectivity at 60-120 mm (left hemisphere). Only the left precuneus cluster remained significant in the global (“all”) condition, where rFCS was computed without distance constraints. Notably, the short-range somatosensory deficit was more pronounced in the right hemisphere, as reflected by larger cluster size and a higher peak statistic (see Table 3). The corrected cluster in the right dorsal S1 at 15-30 mm remained significant even without z-score standardization (aFCS; Supplementary Fig. S1). In sum, distance was a determinant factor for cluster occurrence and direction of effect.

**Figure 1.**
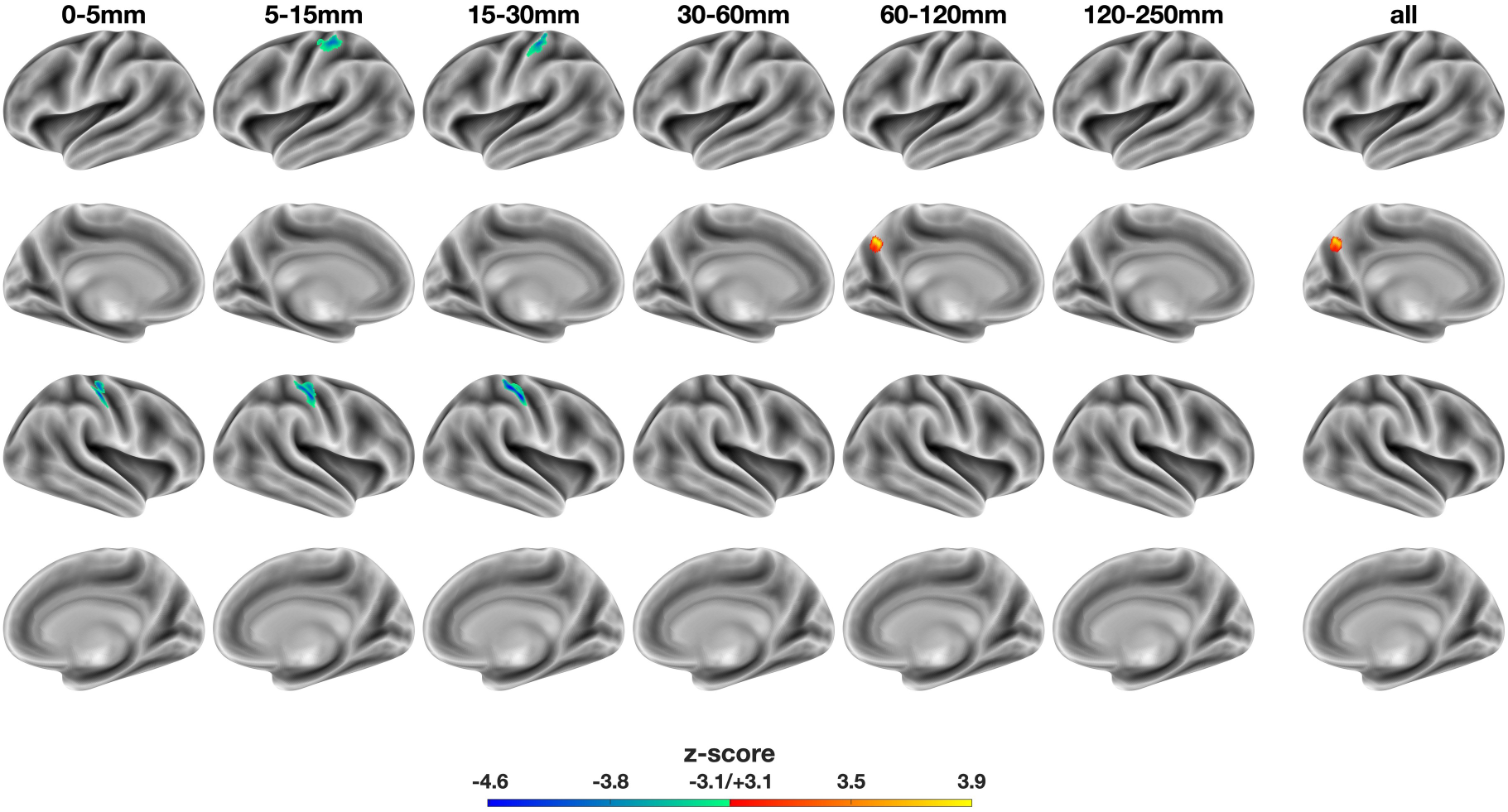
Voxel-wise, distance-dependent differences in relative functional connectivity strength (rFCS): SSD - HC. The colorbar represents z-score, with warm and cold colors indicating increased and decreased rFCS in SSD, respectively. Column labels denote distance ranges used to compute voxel-wise rFCS. The rightmost “all” column shows significant differences in global rFCS (i.e. non-distance-dependent). SSD: schizophrenia spectrum disorder. HC: healthy controls. Notably, the bilateral cluster in dorsal S1 is only significant at short ranges (5-30 mm) and was not identified in the global rFCS (“all”).

**Table 3:**
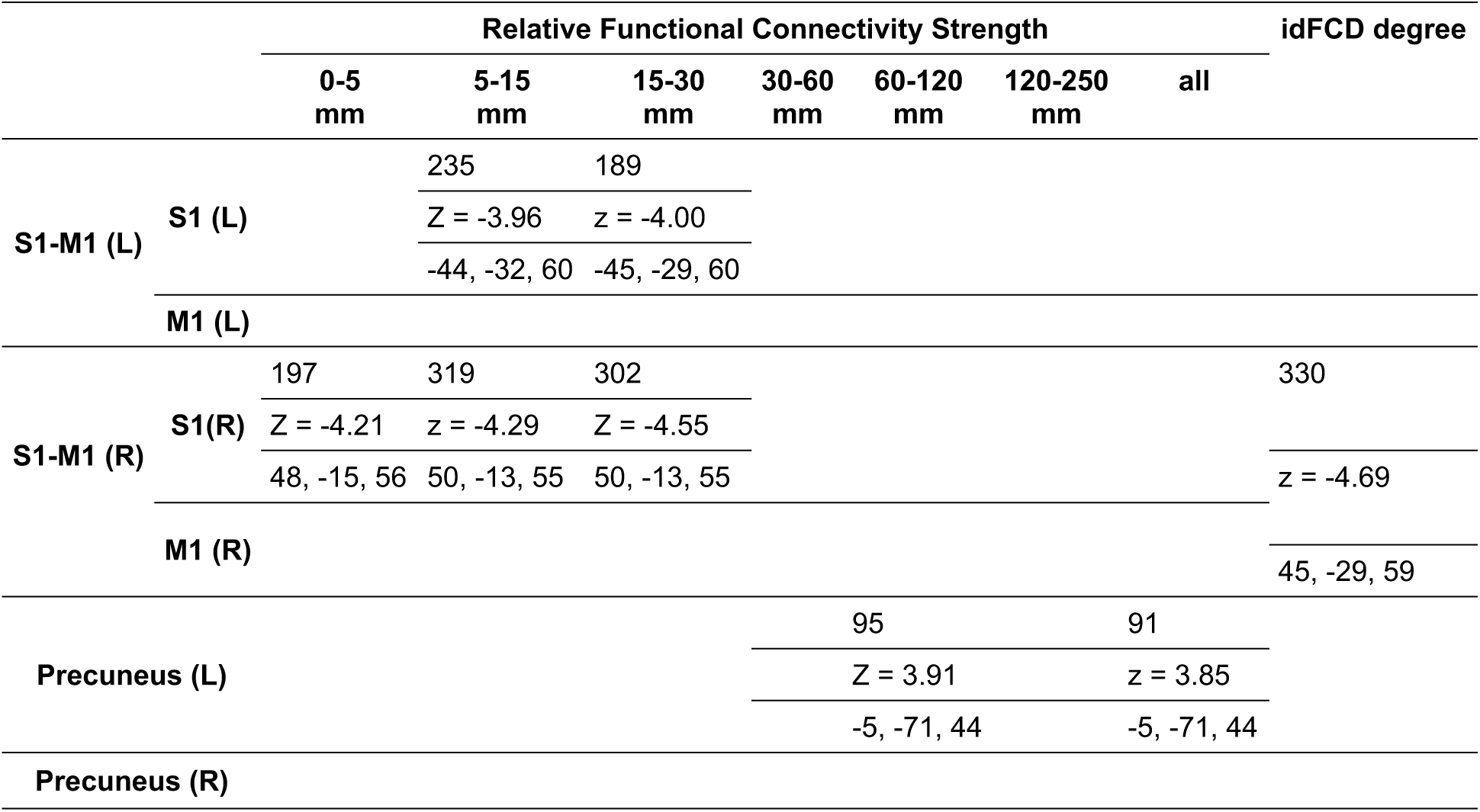
Cluster size (amount of vertices), peak z-score and peak MNI152 coordinates (x, y, z).

#### 3.2.2 FCS across macroscale network hierarchy

Primary network connectivity is reduced in schizophrenia patients, whereas transmodal networks show divergent patterns in rFCS and aFCS. While the distance-dependent analysis identified significant clusters of altered rFCS, examining the unthresholded statistical z-map (Fig. 2A) suggested a broader pattern: schizophrenia patients showed reduced connectivity predominantly in primary cortical areas and increased connectivity in higher-order transmodal networks. To systematically assess how macroscale cortical organization relates to functional connectivity alterations in schizophrenia, we grouped and averaged intra-subject FCS values into three hierarchical levels for each distance range (Fig. 2B).

**Figure 2.**
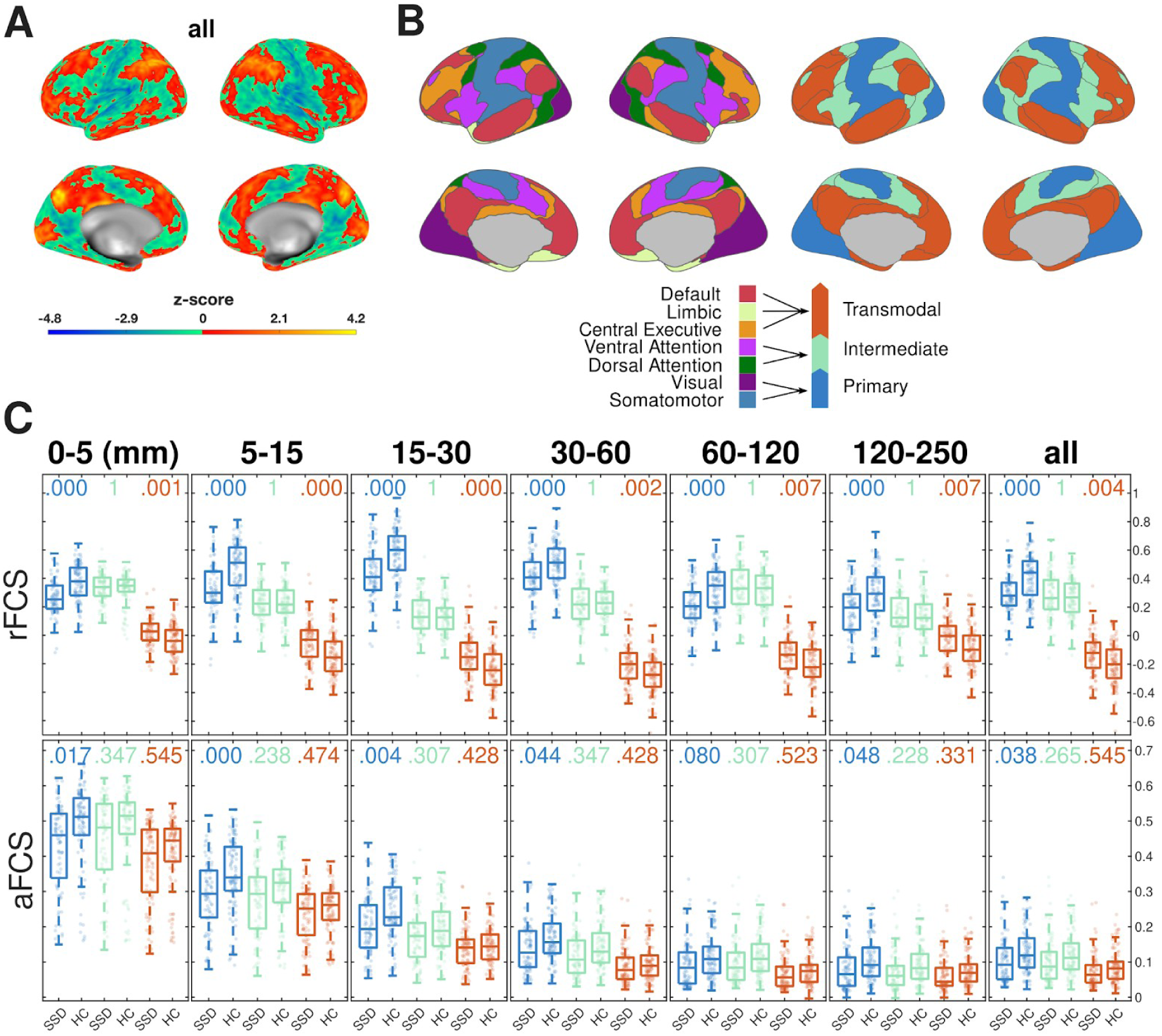
FCS differences grouped by macroscale brain organization. **(A)** Statistical z-map of SSD-HC differences from Fig. 1, showing rFCS alterations before applying the cluster-forming threshold. **(B)** (Left) Parcellation of seven resting-state networks as proposed by Thomas Yeo et al. (2011). (Right) These networks were grouped into a three-level macroscale hierarchy of functional cortical organization according to the principal gradient identified by Margulies et al. (2016): Primary networks (blue), intermediate networks (green), and transmodal networks (red). **(C)** Pair-wise comparisons of mean distance-dependent FCS across the three network-hierarchy levels defined in panel B. Boxplots show the distribution of mean FCS values per group, with individual-level means underlaid as scatter points. P-values for two-tailed tests of mean differences are shown above each boxplot pair, estimated from 10,000 random SSD ⟺ HC label permutations and corrected for FWE using the Holm-Bonferroni method. SSD: schizophrenia spectrum disorder. HC: healthy controls

A three-way LMM controlling for sex, motion confounds, and data set random effects was conducted to assess the independent and interactive effects of network hierarchy (primary, intermediate, and transmodal), distance (six spatial scales + “all”), and diagnosis (SSD vs. HC) on both rFCS and aFCS. Significant main effects were found for network hierarchy (rFCS: F_n_ = 20.74, p<0.001; aFCS: F_n_ = 93.94, p<0.001) and distance (rFCS: F_d_=649.95, p= p<0.001; aFCS: F_d_=62.62, p= p<0.001), indicating that both cortical hierarchy and spatial scale strongly influence functional connectivity.

The main effect of diagnosis was weaker (rFCS: F_g_=3.94, p=0.047; aFCS: F_g_=4.16, p=0.041). However, strong interaction effects were observed between diagnosis and network-hierarchy level (rFCS: F_ng_=53.53, p<0.001; aFCS: F_ng_=33.13, p<0.001), diagnosis and distance (rFCS: F_dg_=5.28, p<0.001; aFCS: F_dg_=31.83, p<0.001), and all three factors combined (rFCS: F_dng_=40.24, p<0.001; aFCS: F_dng_=102.14, p<0.001), suggesting that functional connectivity alterations in schizophrenia depend on cortical macroscale organization including both functional hierarchy and spatial scale. Additionally, network hierarchy interacted with distance, regardless of diagnostic group (rFCS: F_nd_=187.61, p<0.001; aFCS: F_nd_=233.18, p<0.001), which points to distinct connectivity profiles across primary, intermediate, and transmodal networks.

Post-hoc comparisons of pairwise SSD-HC differences within each distance range and network-hierarchy level revealed significant reductions in both rFCS and aFCS within primary cortical networks (comprised of occipital, primary somatomotor, posterior insular, and auditory cortices) (Fig. 2C). Specifically, schizophrenia patients exhibited lower rFCS across all scales and lower aFCS at shorter ranges (≤ 30 mm). In contrast, rFCS was increased in transmodal networks (central executive, limbic, default mode) across all scales, but aFCS values were slightly lower in SSD compared to HC, though not significantly so. No significant differences in either rFCS or aFCS were observed for intermediate networks. FWE-corrected p-values for all comparisons are reported in Fig. 2C.

Taken together, these findings confirm that functional connectivity alterations in schizophrenia are dependent on both spatial scale and hierarchical network organization, with primary networks showing consistent deficits and transmodal networks exhibiting a divergence between relative and absolute connectivity.

### 3.3 idFCD degree and T1/T2 integrity

Schizophrenia patients exhibit inferred cortical microstructural alterations that colocalize only with short-range functional connectivity deficits. Thus far, our results indicate decreased short-range functional connectivity within primary networks (Fig. 2) and particularly in the dorsal primary somatosensory cortex (Fig. 1 and Fig. S1). This raises the question of whether these functional abnormalities are accompanied by corresponding structural changes. Because short-range cortico-cortical structural connections are difficult to detect using white-matter tractography, we employed two indirect measures of cortical structural connectivity: the functionally derived tpFCP and the T1/T2 ratio—a marker of gray matter integrity (see Materials and methods). A statistically significant cluster of reduced idFCD was identified in the right primary sensorimotor cortex in patients compared to controls, affecting dorsal portions of both S1 and M1 (Fig 3A, Table 3). This idFCD deficit likely reflects reduced anatomical connectivity in these regions, as discussed in the methods. A corresponding but non-significant cluster was also observed in the left hemisphere, along with bilateral clusters in the early visual cortex (calcarine and lateral occipital), though these did not survive FWE correction (Fig 3B).

**Figure 3.**
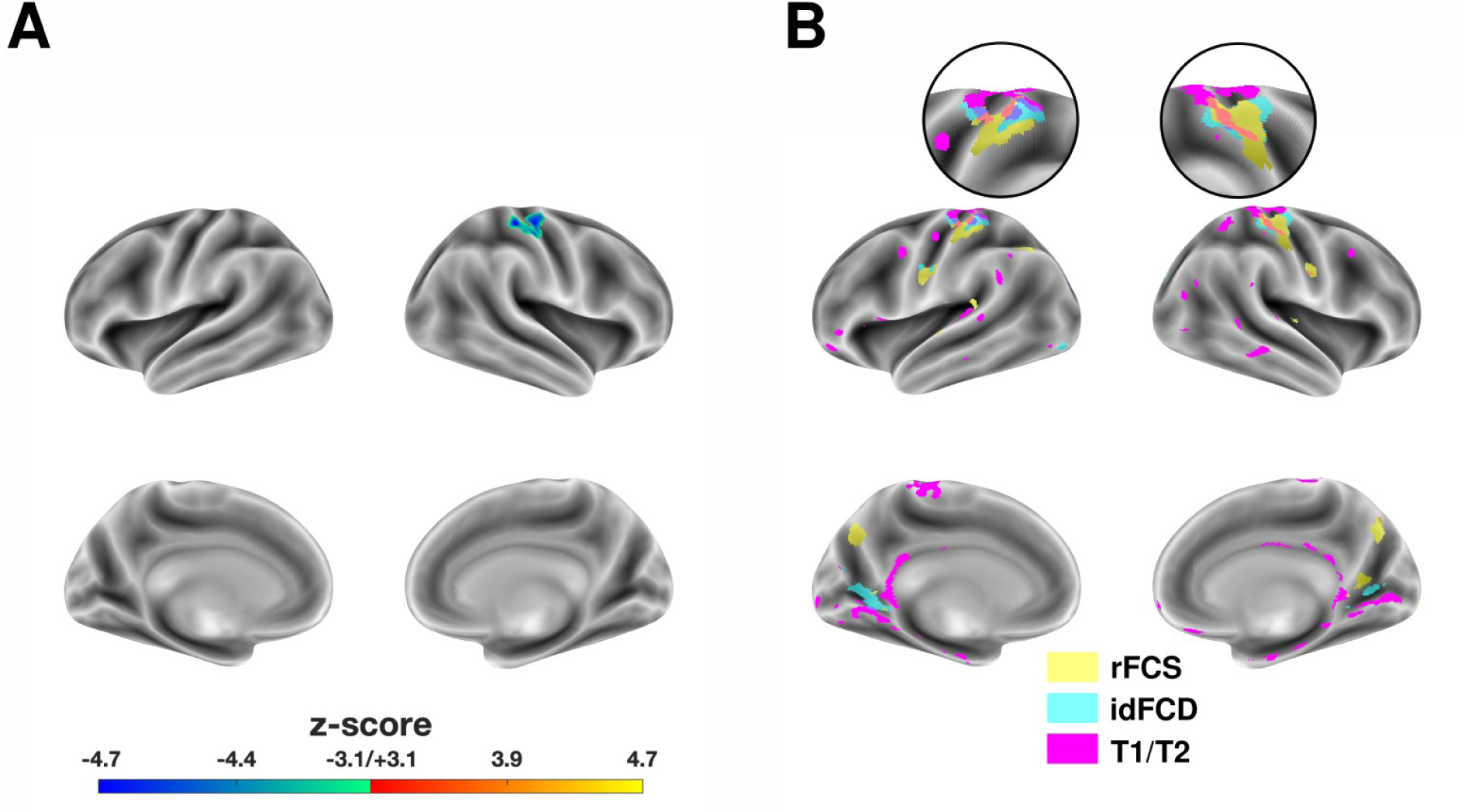
Anatomical correspondence between functional and inferred structural alterations in SSD compared to HCs. **(A)** Right dorsal somatomotor cluster showing significantly decreased idFCD in schizophrenia patients relative to healthy controls (SSD - HC). **(B)** Anatomical colocalization of uncorrected clusters for rFCS (yellow; clusters from all distance ranges were merged), idFCD (cyan), and T1/T2 (magenta). Magnified insets highlight co-localized clusters in the dorsal S1 in each hemisphere, with no other areas showing overlap.

No significant clusters were detected in cortical T1/T2 integrity (as measured by the T1/T2 ratio) after FWE correction. However, uncorrected statistical maps revealed numerous smaller clusters predominantly along the medial walls of both occipital lobes and the dorsal somatomotor cortices near the central sulcus. The latter bilaterally coincided with FCS and idFCD clusters of decreased connectivity (Fig 3B, Table 3). No other overlapping regions were found. The spatial colocalization of functional and inferred structural abnormalities across FCS, idFCD, and T1/T2 ratio based on uncorrected clusters is summarized in Fig 3B.

Inferred structural and functional alterations correlate more strongly at short-ranges in primary networks. Beyond anatomical colocalization, we quantified the spatial covariance between abnormalities in inferred structural measures (idFCD and T1/T2) and rFCS across cortical vertices as a function of network hierarchy. Comparing SSD-HC differences (z-map values) between ΔidFCD and ΔT1/T2 revealed a moderate positive correlation in primary networks, a weaker correlation in intermediate networks, and a negligible correlation in transmodal networks (Fig. 4A) (r_Primary_ = 0.297, r_Intermediate_ = 0.222, r_Transmodal_ = 0.018). At shorter distances, pairwise comparisons between ΔrFCS and ΔT1/T2 followed a similar trend, with the strongest correlations in primary networks, weaker correlations in intermediate networks, and weak negative correlations in transmodal networks. For example, at the 5-15 mm scale (Fig. 4A), ΔrFCS vs. ΔT1/T2 yielded r_Primary_ = 0.293, r_Intermediate_ = 0.125, r_Transmodal_ = -0.099. This hierarchical ordering was consistent for distances below 30 mm but collapsed at longer distances. For the longest distance range, correlations uniformly increased again, which may reflect longer-range white-matter alterations captured by the T1/T2 ratio (Fig. 4B). Correlations between ΔrFCS and ΔidFCD also showed a similar hierarchical pattern at shorter distances, but with stronger coefficients, consistent with their shared dependence on functional connectivity. At 5-15 mm, the correlation coefficients were r_Primary_ = 0.743, r_Intermediate_ = 0.664, r_Transmodal_ = 0.474, with values decreasing as distance increased.

**Figure 4.**
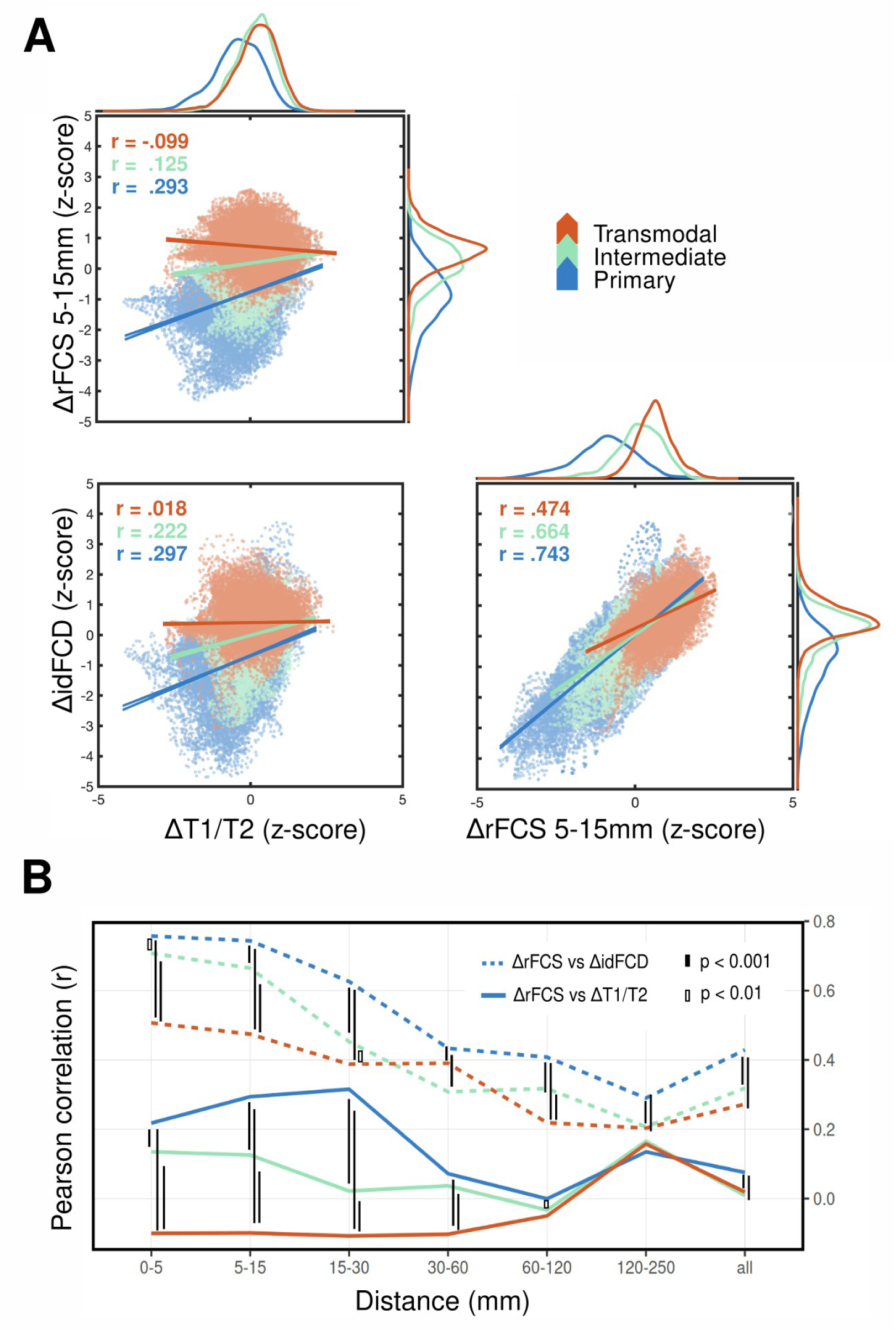
Statistical correlations between functional and inferred structural alterations in SSD compared to HCs. **(A)** Example pairwise correlations of SSD-HC differences in functional and inferred structural measures. Each sub-panel compares two difference measures (ΔrFCS at 5-15 mm, ΔidFCD, and ΔT1/T2) in units of z-score deviations, obtained from the previous vertex-wise statistical tests (SSD vs. HC). Each point in the scatter plot thus represents a vertex-wise SSD-HC z-score pair. Linear regression trends and Pearson correlation coefficients (r) were computed separately for each network hierarchy level. Vertices belonging to primary, intermediate, and transmodal networks are color-coded in blue, green, and red, respectively. Linear trends are shown with their 95% confidence intervals obtained from simple linear regression models. Correlation in SSD-HC differences between ΔidFCD, and ΔT1/T2 are stronger for primary networks than for intermediate and transmodal networks. This is also the case for correlations of ΔidFCD, and ΔT1/T2 with ΔrFCS at 5-15 mm, but not at all distances as shown in (B). **(B)** Variation of correlation strength across distance ranges and network-hierarchy. Solid lines indicate correlations between ΔrFCS and ΔT1/T2, while dashed lines indicate correlations between ΔrFCS and ΔidFCD. FWE-corrected significant differences between hierarchy levels at a given distance range are indicated by black horizontal connector lines of varying fill, corresponding to different p-value thresholds (see legend). Also see Fig. S2 for the remaining post-hoc pairwise comparisons among hierarchical levels alone and distance ranges alone.

A two-way ANOVA confirmed statistically significant effects of network hierarchy, distance, and their interaction on correlation coefficients for both ΔrFCS vs. ΔT1/T2 (F_n_ = 1683.79, F_d_ = 226.11, and F_nd_ = 204.29 with p < 0.001 for all) and ΔrFCS vs. ΔidFCD (F_n_ = 844.04, F_d_ = 1014.2, and F_nd_ = 44.80 with p < 0.001 for all). Finally, correlations between the two inferred structural measures also showed a significant main effect of network hierarchy (ΔidFCD vs. ΔT1/T2: F_n_ = 349.14, p < 0.001).

Post-hoc pairwise comparisons confirmed the qualitative trends observed in Fig. 4B. That is, the distributions of cross-product deviations—used to compute Pearson correlations—differed significantly across the network hierarchy (see Materials and methods). This effect was most pronounced for distances below 30 mm, while differences disappeared at longer distances for ΔrFCS vs. ΔT1/T2.

Together, these vertex-wise and macroscale results indicate abnormal connectivity in primary cortical networks in schizophrenia, with the most pronounced deficits in the dorsal somatomotor cortex. Moreover, microstructural changes relative to healthy controls reinforce the relevance of short-range functional connectivity disruption in these regions.

### 3.4 Correlation between rFCS and psychiatric symptoms in the COBRE dataset

To further explore the clinical relevance of the identified connectivity alterations, we examined correlations between average rFCS and PANSS symptom dimensions in the COBRE sample, including both individual subscale scores and principal components. Although none of the symptom correlations survived correction for multiple comparisons, average rFCS within clusters of reduced connectivity in the right dorsal primary somatosensory cortex (S1) showed negative correlations with negative PANSS symptoms (5–15 mm: r = –0.31, p = 0.032; 15–30 mm: r = –0.39, p = 0.017, uncorrected, Fig. S3A), consistent with the direction of group-level differences. The left hemisphere homologues correlated negatively with age. We performed PCA on the PANSS scores to better characterize symptom structure (Fig. S3E,F). The first component coincided with age (explaining 81% of variance). The second principal component (5% of the variance) built on positive symptoms such as delusions, hallucinatory behavior and suspiciousness; as well as some general symptoms including anxiety, depression, guilt feelings and unusual thought content. The third principal component (4% of the variance), which heavily loaded on expressive and motor-related negative symptoms, including poor rapport, blunted affect, and motor retardation, also negatively correlated with the right S1 cluster rFCS at 15–30 mm (r = –0.35, p = 0.02, uncorrected). This component further showed negative correlations with primary network rFCS at longer distances (>60 mm and all distances combined) and positive correlation with transmodal network rFCS at 15–30 mm (r = 0.32, p = 0.03, uncorrected) (see Fig. S3B,D for complete distance-by-hierarchy analysis). While preliminary, these trends suggest a potential link between local somatomotor hypoconnectivity and both negative and motor-expressive symptoms in schizophrenia.

## 4. Discussion

To capture connectivity alterations in schizophrenia that may be undetectable using traditional region-based analyses, we employed a distance-dependent, vertex-wise approach to measuring FCS. This method complements prior work that has primarily focused on connectivity between larger, predefined brain regions and allows for the detection of localized, spatially constrained dysconnectivity that may reflect underlying microstructural abnormalities, which are hard to detect using structural-connectivity neuroimaging, such as diffusion-weighted MRI.

We found that short-range FCS was significantly reduced in the dorsal primary somatosensory cortex (S1) in schizophrenia patients. This result was not evident in the global, distance-agnostic FCS analysis, highlighting the value of incorporating explicit distance information into FCS assessment. Moreover, these short-range FC deficits colocalized with two independent indirect measures of cortical microstructure: idFCD and T1/T2 ratio. Both measures revealed reduced structural integrity in overlapping regions of the somatomotor cortex, consistent with localized structural disruption as a plausible explanation for the decreased short-range FCS. The idFCD cluster in the right dorsal S1/M1 survived cluster correction, while T1/T2 abnormalities, though not significant after correction, showed similar spatial patterns in uncorrected maps.

In contrast to the short-range reductions, we observed increased longer-range FCS in the dorsoposterior precuneus, particularly in the 60–120 mm range. This effect was significant for rFCS and evident, though not statistically significant after correction, for aFCS. In addition, this long-range increase did not spatially overlap with abnormalities in either idFCD or T1/T2 ratio, suggesting it may reflect network-level reorganization in transmodal areas. However, the absence of detectable local microstructural differences does not rule out the possibility of underlying anatomical disruption, specifically, alterations in white-matter tracts between regions, which are more directly assessed using diffusion-weighted imaging.

Previous studies have explored spatially constrained forms of FC in SSD, for instance by setting a single distance threshold (typically 75 mm) before measuring the average (FCS) or thresholded binary sum (functional connectivity density, FCD) of correlations between a given time series and the rest of the brain (X. Wang et al. 2014; W. Guo et al. 2015; 2017; Miao et al. 2020). Reduced short-range FCS (< 75 mm) has been reported in primary sensory and motor regions including the postcentral gyrus, supplementary motor area, insula, and visual cortex (X. Wang et al. 2014; Miao et al. 2020), consistent with the deficits we observed in dorsal somatosensory cortex. In contrast, increases in both short- and long-range FCS have been reported in default mode and frontal regions, including the medial prefrontal cortex, inferior parietal lobule, and posterior cingulate cortex (W. Guo et al. 2015; 2017). However, these studies measured anatomical distance as the Euclidean distance between MNI coordinates and included interhemispheric connections in the “short-range” (< 75 mm) category. By contrast, our method measured geodesic distances on the inflated cortical surface and treated hemispheres separately, thereby offering a more anatomically grounded assessment of distance-dependent cortico-cortical connectivity.

Another approach to evaluating local connectivity alterations in schizophrenia uses higher-order Kendall correlation (regional homogeneity, ReHo) among strictly adjacent voxels (C. Zhao et al. 2018; Yu et al. 2021; Cai et al. 2022). Cai et al. (2022), which included the COBRE and NMorphCH datasets analyzed here, reported decreased ReHo in the bilateral postcentral and precentral gyri and increased ReHo in the bilateral medial superior frontal gyrus, although the latter was not significant in the NMorphCH dataset. While these findings align with our results, our distance-dependent analysis further revealed that local FCS deficits in somatomotor regions are not limited to nearest-neighbor scales, but extend up to 30 mm, indicating a more spatially extensive disruption of local functional organization. Taken together, findings from both regional FCS studies and ReHo analysis converge on a pattern of reduced short-range connectivity in primary sensory-motor regions and connectivity increases at multiple spatial scales in transmodal networks in schizophrenia.

Consistent with these observations, our hierarchical analysis revealed a similar dissociation, with robust FCS reductions in primary networks across all spatial scales, particularly for short-range connections (≤30 mm), and increased rFCS in transmodal networks regardless of distance. Notably, these increases were not accompanied by corresponding overall increases in aFCS, suggesting a relative redistribution of connectivity rather than uniform hyperconnectivity. However, the general spatial pattern of increased and decreased connectivity within transmodal regions (Fig. 2A) was preserved using aFCS. The increase in rFCS in patients vs. controls may thus be driven by more specific transmodal regions such as the precuneus identified in the voxel-wise analysis and the inferior parietal cortex (W. Guo et al. 2017; L. Guo et al. 2023). No group differences were found for intermediate networks. Correlations between SSD–HC differences in rFCS and inferred structural measures (T1/T2 and idFCD) were strongest for primary networks at short-range distance (≤30 mm), whereas differences in transmodal rFCS and T1/T2 showed weak, negative correlations at shorter ranges. While differential functional connectivity alterations occur in primary and transmodal regions, only reductions in primary regions show consistent colocalization with local microstructural differences. Because earlier studies have primarily relied on regional or whole-brain voxel-wise analyses and diffusion-weighted imaging, which are more sensitive to long-range white-matter tracts and often highlight transmodal networks such as the DMN, this short-range functional and local microstructural dysconnectivity in primary networks may have been systematically overlooked. Our results thus provide rare evidence of a direct structure–function relationship in schizophrenia within primary sensory areas.

Moreover, identified group differences were concentrated in regions that, in healthy individuals, exhibit both high local functional connectivity and high intracortical myelination— features that characterize the unimodal end of the principal cortical gradient from primary to transmodal networks (Margulies et al. 2016; Huntenburg et al. 2017). While our approach is more sensitive to local disruptions, the fact that significant group differences emerged specifically in highly myelinated, locally connected cortical regions suggests that these regions may be particularly vulnerable to disease-related dysconnectivity—an effect that may be missed in studies emphasizing long-range or global alterations. Notably, the principal gradient identified by Margulies et al. (2016) also aligns with increasing geodesic distance along the cortical surface: transmodal regions are maximally distant from unimodal sensorimotor areas. Complementing prior evidence of transmodal dysconnectivity, our findings suggest that schizophrenia involves distinct modes of dysconnectivity that align with both spatial and hierarchical gradients of cortical organization: short-range reductions in primary sensory areas that colocalize with microstructural abnormalities, and longer-range increases in transmodal regions that appear structurally decoupled at the local level.

Two recent studies have directly examined alterations in cortical gradients in schizophrenia by projecting patient data onto the principal connectivity gradient originally defined by Margulies et al. (2016). Dong et al. (2021) reported a compression of the principal gradient in schizophrenia, characterized by reduced segregation between sensorimotor and transmodal areas, consistent with both hypoconnectivity within primary sensory regions and hyperconnectivity with frontoparietal areas. In contrast, Holmes et al. (2023) found no significant disruption of the principal gradient in early psychosis or schizophrenia but reported altered organization along a secondary visual–sensorimotor gradient in patients with established illness. Although their conclusions regarding the principal gradient differ, both studies describe alterations that align with our findings. Rather than projecting onto gradient space, we assessed distance-dependent FCS stratified by primary, intermediate, and transmodal networks. In line with Dong et al. (2021), we observed differential functional alterations across the cortical hierarchy, with reduced FCS in primary networks and increased rFCS in transmodal regions. In line with Holmes et al. (2023), short-range alterations were most prominent in somatomotor areas, which may underlie the observed reduction in the secondary visual-sensorimotor gradient.

In a recent large-sample study, Schleifer et al. (2025) investigated regional alterations in local and global connectivity in individuals with 22q11.2 deletion syndrome, a genetic condition associated with psychosis, and adolescents at clinical high risk for psychosis (CHR), and mapped the results to published biological pathways, including Margulies’ principal gradient and a T1w/T2w ratio map. Notably, only bilateral somatomotor cortices showed decreased connectivity across both groups: local and global connectivity reductions in the 22q11.2 group and local reductions only in the CHR group compared to controls. Additional local connectivity decreases in frontal and parietal regions were observed in the 22q11.2 group but not in CHR. Local connectivity differences between CHR and controls were positively correlated with the principal gradient map, consistent with our findings in schizophrenia patients, while a negative correlation with the gradient was observed in the 22q11.2 group. No significant correlations with T1w/T2w ratio were found in the left hemisphere, and data for the right hemisphere were unavailable. Supporting a link between local functional dysconnectivity and intracortical myelination in schizophrenia, a separate large-sample study by Jørgensen et al., (2016) reported increased cortical gray/white matter contrast in primary sensory and motor regions, particularly the post- and precentral gyri, an effect interpreted as indicative of reduced intracortical myelination in these areas.

Several limitations of this study should be acknowledged. First, our analyses were conducted in a cross-sectional sample of individuals with chronic schizophrenia receiving antipsychotic medication. Although antipsychotic effects on functional connectivity cannot be excluded, converging evidence from at-risk populations suggests that our findings are not solely driven by medication or chronic illness (Schleifer et al. 2025). Nonetheless, replication of our distance-dependent analysis in early-stage, medication-naive cohorts is needed to confirm the developmental and treatment-independent nature of the observed effects.

Second, the measures used to infer microstructural alterations are indirect. The idFCD measure reflects putative anatomical connections based on high functional connectivity values, but this approach may miss weaker or distributed effects due to a higher rate of false negatives. The T1w/T2w ratio, while widely used as a proxy for intracortical myelination, remains a debated measure with respect to its specificity (Mühlau 2022; Sandrone et al. 2023). Importantly, these two measures are methodologically independent, yet both converged on disruptions within the somatosensory cortex in the cluster-based analysis and showed the strongest correspondence with short-range FCS reductions in primary networks in the hierarchical analysis, lending robustness to the structural interpretation of our functional findings.

Third, our analysis focused exclusively on cortico-cortical connectivity using a surface-based approach, which allowed for anatomically precise geodesic distance measurements but excluded subcortical regions from consideration. Consequently, we did not assess functional or structural connectivity involving subcortical structures, particularly the thalamus, which plays a central role in cortico-thalamo-cortical communication and has been consistently implicated in schizophrenia (Giraldo-Chica and Woodward 2017). Prior studies have reported thalamic hypoconnectivity with prefrontal regions and hyperconnectivity with sensory and motor cortices in schizophrenia (Woodward et al. 2012; Anticevic et al. 2014). This thalamic hyperconnectivity has moreover been associated with decreased FC within sensorimotor networks (Kaufmann et al. 2015; Baran et al. 2019). Notably, Baran et al. (2019) further showed that thalamocortical hyperconnectivity with somatosensory and motor cortices was inversely correlated with reduced sleep spindle activity, a physiologically and clinically relevant marker of thalamocortical dysfunction in schizophrenia (Ferrarelli and Tononi 2017; Kozhemiako et al. 2025).

Finally, our goal was to investigate FCS alterations in a distance-dependent manner using vertex-wise data without relying on predefined regions, thereby complementing standard region-based FC approaches. In parallel, we evaluated two independent proxy measures of local microstructure to augment more commonly used diffusion-weighted imaging measures. Because the number of vertices considered in the FCS average increases with distance, our approach is inherently more sensitive to short-range alterations. Nevertheless, we identified a significant cluster of increased rFCS in the 60-120 mm range, indicating that longer-range changes can also be detected. Importantly, these methods provide complementary insights to interareal FC analyses by revealing short-range dysconnectivity that may not be captured in global measures. While our results point to a convergence between short-range functional and structural alterations in primary networks and the somatomotor cortex in particular, future work employing interventional or model-based methods such as effective connectivity analysis will be essential for elucidating the directionality and causal relationships between these observed network disruptions.

In conclusion, this study provides novel evidence for spatially and hierarchically specific dysconnectivity in schizophrenia, using a distance-dependent, vertex-wise approach to functional connectivity analysis. We identify robust reductions in short-range connectivity within the primary somatosensory cortex that colocalize with two independent measures of inferred microstructural disruption. These findings suggest a direct structure–function relationship that has been overlooked by traditional region-based and diffusion-weighted imaging studies. In contrast, increased connectivity in transmodal networks was not associated with local structural differences, potentially reflecting alterations in large-scale white matter pathways. Together, these findings highlight two distinct patterns of cortical dysconnectivity and underscore the value of considering spatial scale and cortical hierarchy in future research.

## Data Availability

All data produced are available online at https://schizconnect.org

## Supplement

**Figure S1.**
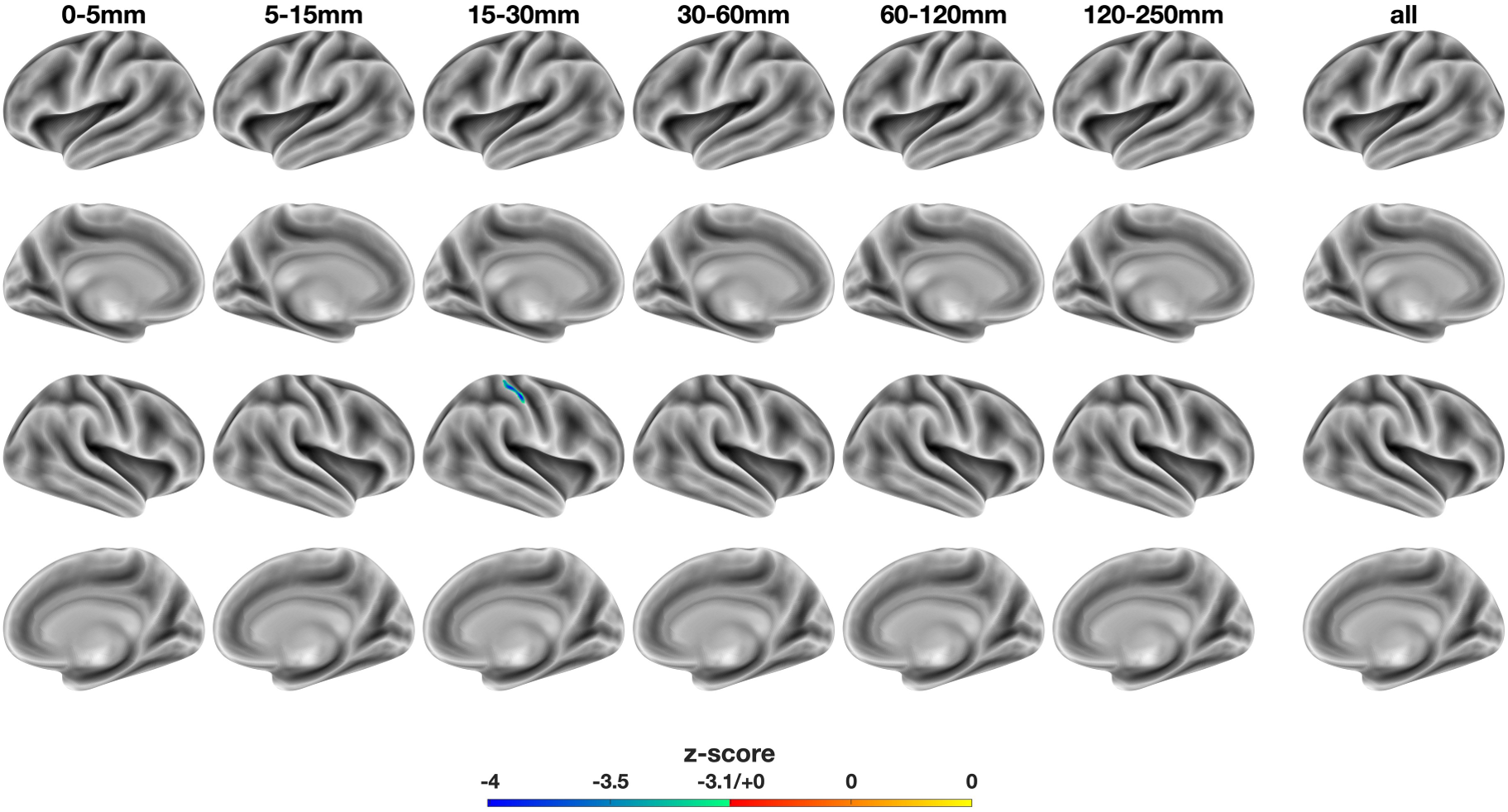
Voxel-wise, distance-dependent differences in absolute functional connectivity strength (aFCS): SSD - HC. The colorbar represents z-score, with warm and cold colors indicating increased and decreased aFCS in SSD, respectively. Column labels denote distance ranges used to compute voxel-wise aFCS. The rightmost “all” column shows significant differences in global aFCS (i.e. non-distance-dependent). SSD: schizophrenia spectrum disorder. HC: healthy controls. Only the right dorsal S1 cluster at 15-30 mm remains significant in the aFCS analysis.

**Figure S2.**
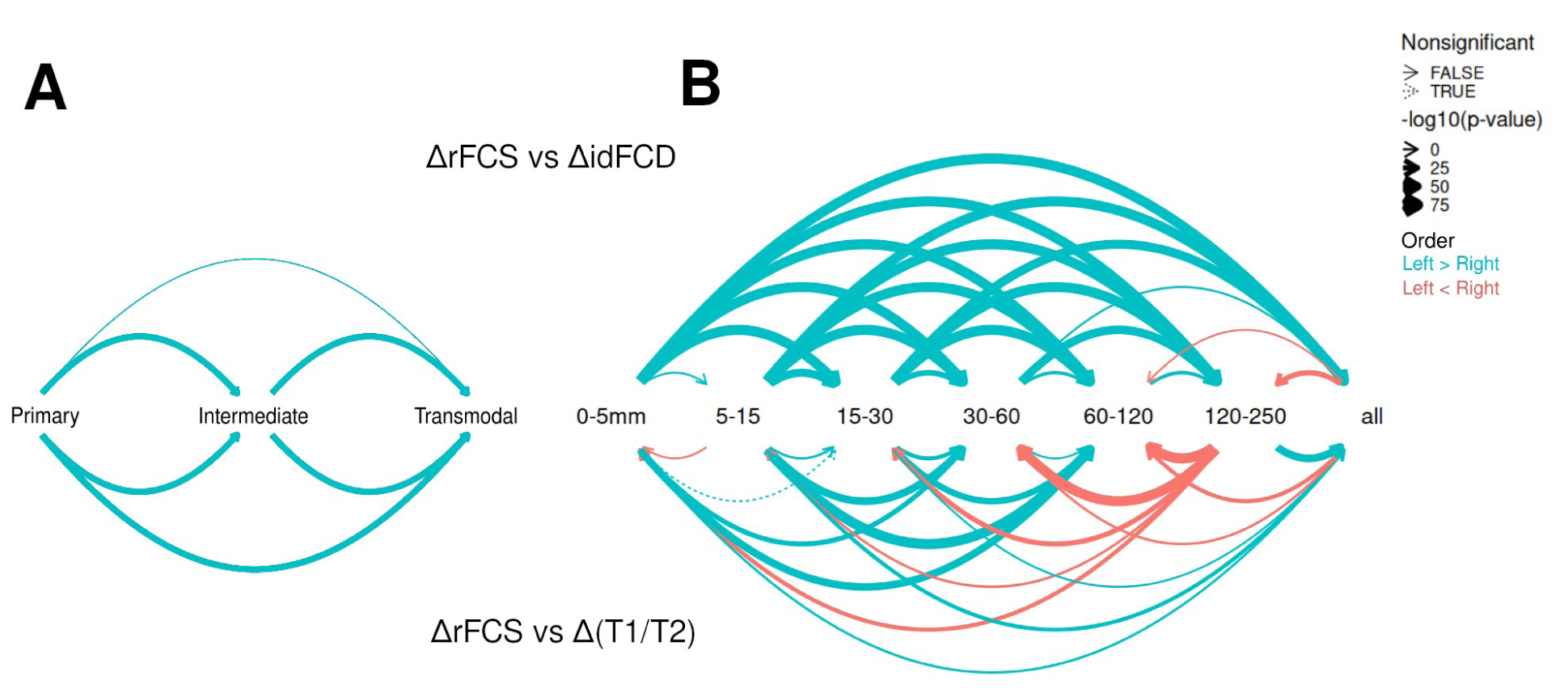
Statistical comparisons between correlations in functional and inferred structural alterations in SSD compared to HCs (continued from Fig 4B). Remaining post-hoc pairwise comparisons among groups of correlations of SSD-HC differences in functional and inferred structural measures. (A) Comparisons among levels when grouping by network hierarchy alone. (B) Comparisons among levels when grouping by distance alone. In both cases, graph edges in the upper half represent comparisons of correlations between ΔrFCS and ΔidFCD. Lower half represents ΔrFCS and Δ(T1/T2). The direction of the effect (group with higher mean cross-product deviations) is given by arrow direction and color, with cyan standing for stronger correlations to the left of the plot. Smaller p-values (FWE-corrected) translate into thicker arrows. In general, primary networks display stronger correlations between functional and inferred structural alterations, which in turn display stronger correlations than transmodal networks. Similarly, correlations become weaker as more distant alterations in rFCS are considered.

**Figure S3.**
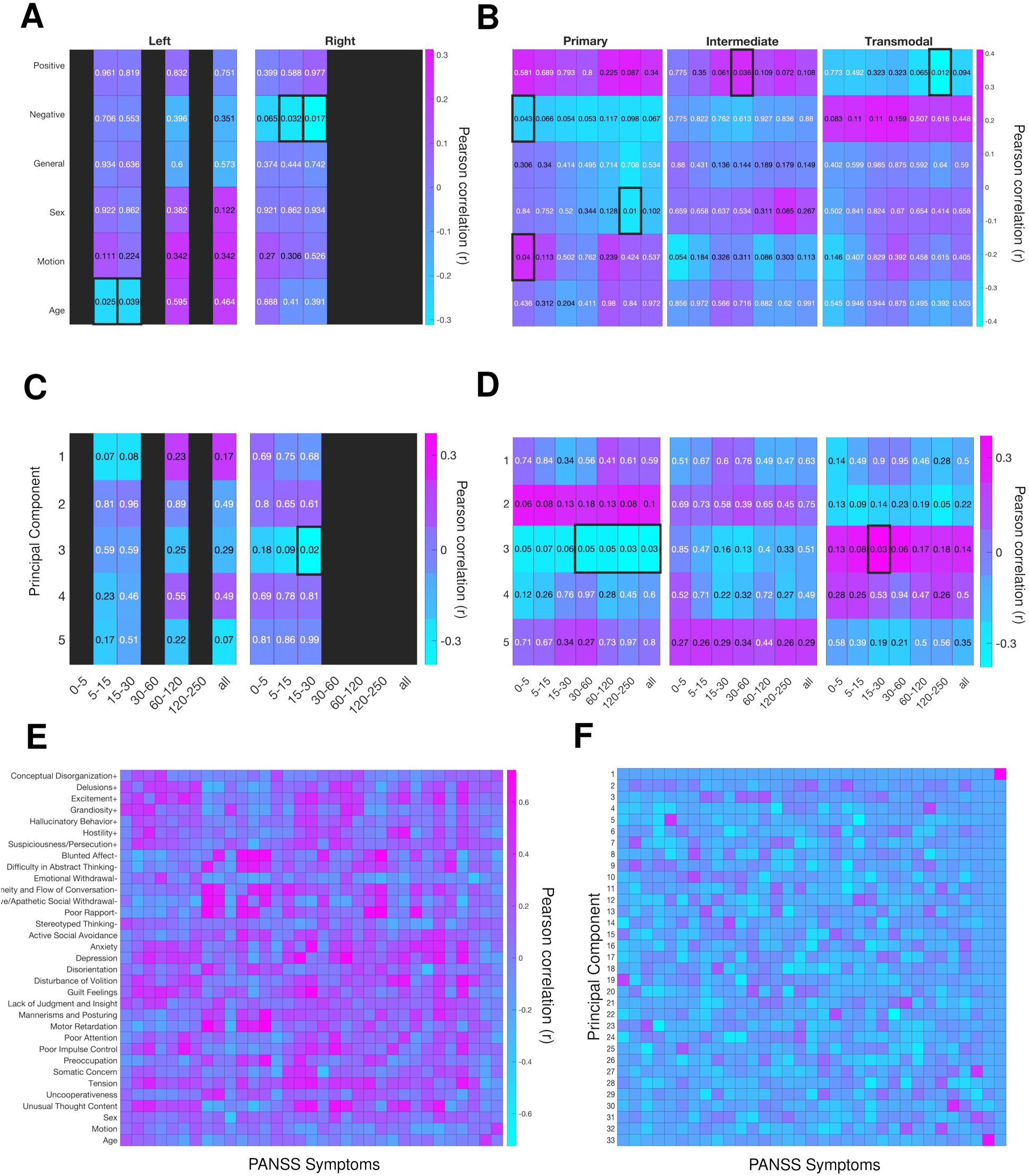
Correlation between rFCS and psychiatric symptoms in the COBRE dataset. **(A)** Pearson correlations of PANSS subscale scores (positive, negative, general, plus confound variables) against individual-level average rFCS from significant clusters in Fig. 1. Number at each cell is the (uncorrected) p-value corresponding to the parameter for that symptom, according to a multiple linear regression fit of rFCS. Colorbar represents Pearson correlation, with pink for positive and blue for negative associations, respectively. **(B)** Same as (A) but correlating against average rFCS from hierarchy levels (primary, intermediate, and transmodal networks). **(C)** Same as (A) but using the principal components from (F) (only first 5 PC shown here), instead of original PANSS subscale scores and confounds. **(D)** Same as (C) but correlating against average rFCS from hierarchy levels (primary, intermediate, and transmodal networks). **(E)** Correlation matrix across patient scores for all 30 symptoms in the Positive and Negative Syndrome Scale (PANSS). **(F)** Projection of original PANSS scores on the components found by PCA.

